# The anti-vaccination infodemic on social media: a behavioral analysis

**DOI:** 10.1101/2020.12.07.20223370

**Authors:** Federico Germani, Nikola Biller-Andorno

## Abstract

Vaccinations are without doubt one of the greatest achievements of modern medicine, and there is hope that they can constitute a solution to halt the ongoing COVID-19 pandemic. However, the anti-vaccination movement is currently on the rise, spreading online misinformation about vaccine safety and causing a worrying reduction in vaccination rates worldwide. In this historical time, it is imperative to understand the reasons of vaccine hesitancy, and to find effective strategies to dismantle the rhetoric of anti-vaccination supporters. For this reason, we analyzed the behavior of anti-vaccination supporters on the platform Twitter. Here we identify that anti-vaccination supporters, in comparison to pro-vaccination supporters, share conspiracy theories and make use of emotional language. We demonstrate that anti-vaccination supporters are more engaged in discussions on Twitter and share their contents from a pull of strong influencers. We show that the movement’s success relies on a strong sense of community, based on the contents produced by a small fraction of profiles, with the community at large serving as a sounding board for anti-vaccination discourse to circulate online. Surprisingly, our data demonstrate that Donald Trump, together with members of his entourage and his closest supporters, are the main drivers of vaccine misinformation on Twitter. Based on these results, we propose to strategically target the anti-vaccination community online through policies that aim at halting the circulation of false information about vaccines. Based on our data, we also propose solutions to improve the communication strategy of health organizations and build a community of engaged influencers that support the dissemination of scientific insights, including issues related to vaccines and their safety.

---

Vaccinations are a great medical achievement of the last century, given their fundamental contribution to lowering the presence of otherwise widespread diseases in the population and thus in greatly reducing mortality. Despite the available evidence and the scientific consensus on the necessity and the safety of vaccines, an anti-vaccination movement has been growing over the past decades *(1)*, with a consequent decline in vaccination rates and the possible resurgence of diseases such as measles *(2)*. This movement, which has gained momentum after the infamous publication of Andrew Wakefield’s study linking vaccines to autism in 1998 *(3)*, has been lately growing its strength, taking advantage of social media as communication channels *(4,5)*. In a postmodern world in which medical expertise is being questioned *(6,7)*, the growing grip of the anti-vaccination movement on the general public is of great concern, especially amidst a global pandemic that could be solved by the development of safe and effective vaccines. Therefore, while we navigate through the COVID-19 pandemic and the concomitant infodemic, the importance of presenting proper information concerning vaccines to the public is of utmost importance.

In order to tackle the vaccination issue, the causes of the success of the anti-vaccination movements need to be carefully analyzed. Until now, it has been shown that vaccination choice is influenced by the belief in alternative medicine, the belief in conspiracy theories, by morality, religion and personal ideology, the emotive appeals or the lack of trust in authorities *(8)*, as well as by the readability and engagement of pro-versus anti-vaccination articles *(9)*. Most studies primarily focus on two aspects, the psychological attitude connected to vaccination choice *(10-12)* and the role of the Internet and in particular social media *(8, 13-18)*. In fact, anti-vaccination supporters find fertile ground in particular on Facebook and Twitter *(17, 19-20)* as these platforms offer a digital space for people to share any kind of content, including science-related or medically sensitive contents, which have the potential to reach a vast audience. Studies have particularly focused on the relevance of the Internet and social media in shaping personal or parental choice about vaccination *(13, 14, 17)*. For instance, parents who decide not to vaccinate their children tend to shape their opinions after having been in contact with online information on the topic *(21)*, and the majority of individuals does not consider the credibility of the source of information *(22-25)* In addition, anti-vaccination profiles and groups online have been shown to generate content that is based on personal experiences and opinions, whereas pro-vaccination groups and institutions have the tendency to quote experts and cite scientific literature when sharing their views online *(9, 23)*. Therefore, the adopted language, the frequency of use of social media, the type of content that is generated, and their emotional appeal, could all constitute factors that determine the success of the anti-vaccination movement online. In order to understand whether these factors are particularly relevant, to identify strategies to decrease the spread of vaccine misinformation online, and to identify potential communication strategies to be used by healthcare organizations and professionals, we decided to quantitatively analyze the online behavior of Twitter users, after having determined whether they support or contrast vaccination programmes.

A recent study has identified US President Donald Trump likely to be the largest driver of the COVID-19 misinformation infodemic *(26)*. This is relevant because fake news, of any kind, have been shown to have affected various democratic votes, including the 2016 US elections and Brexit *(27-29)*. For example, before the 2016 US elections, fake news stories favoring Trump were shared 30 million times on Facebook, against 8 million times for those favoring Clinton *(27)*. For some politicians, social media and fake news, including those concerning vaccines, could therefore be instrumental to hold on power and determining the future course of our global society. In particular, vaccination policies are not excluded from the aspects that could determine the results of the upcoming 2020 US elections, especially in light of the COVID-19 pandemic. In fact, both vaccine hesitancy and political populism are driven by the distrust in expertise and the ideal of a bottom-up society *(30)*, and the political views play an important part in shaping vaccination choice *(31)*.

## Anti-vaccination supporters tweet less, but engage more in discussion

In order to understand whether the success of the anti-vaccination discourse is due to a particularly pronounced activity of anti-vaccination supporters online, we measured the number of Twitter actions on average in a month for each profile belonging to the control, anti-vaccination and pro-vaccination group (Fig. 1A). Control profiles were selected for the use of randomly chosen hashtags (#control). Anti-vaccination users were identified for their use of the #vaccineskill and #vaccinesharm hashtags, which are widely used by the community. Finally, pro-vaccination communicators were identified for their use of the #vaccineswork hashtag (Fig. S1). We defined Twitter actions as the sum of tweets, replies and retweets in a given month (Fig. 1B). As expected, anti-vaccination profiles are the most active on Twitter, with 536 actions per month, compared with an average of 277 actions for the control group and only 144 actions for the pro-vaccination group (Fig. 1C), suggesting the latter is not engaged enough, and highlighting a first pitfall in the pro-vaccine communication strategy online. However, once we calculated the number of tweets per month, we were surprised to learn that anti-vaccination supporters were those tweeting the least (42 tweets per month), when compared with control and pro-vaccination profiles (123 and 93 tweets per month, respectively) (Fig. 1D). This was largely compensated by the engagement of the anti-vaccination group in discussions, be it through replies or retweets. Anti-vaccination profiles replied 13-times more than control and pro-vaccination profiles (Fig. 1E), retweeted 7.4 times more than their pro-vaccination counterparts, and 31.3 times more than control profiles (Fig. 1F). As already pointed out by these data, the anti-vaccination group scored the highest number of retweets per Tweet (Fig. S2), highlighting that the vast majority of anti-vaccination supporters act as an echo chamber for the pool of content generated by a small fraction of users. Behavioral outliers, which were excluded with 0.1% confidence interval (ROUT, Q=0.1%) (data not shown), suggest that a small fraction of users belonging to this group are producing the majority of the content, which is then shared by the community at large. Data also suggest that pro-vaccination individuals and groups are more prone to generate new content and are not very engaged with a broader community with similar interests.

**Figure 1.**
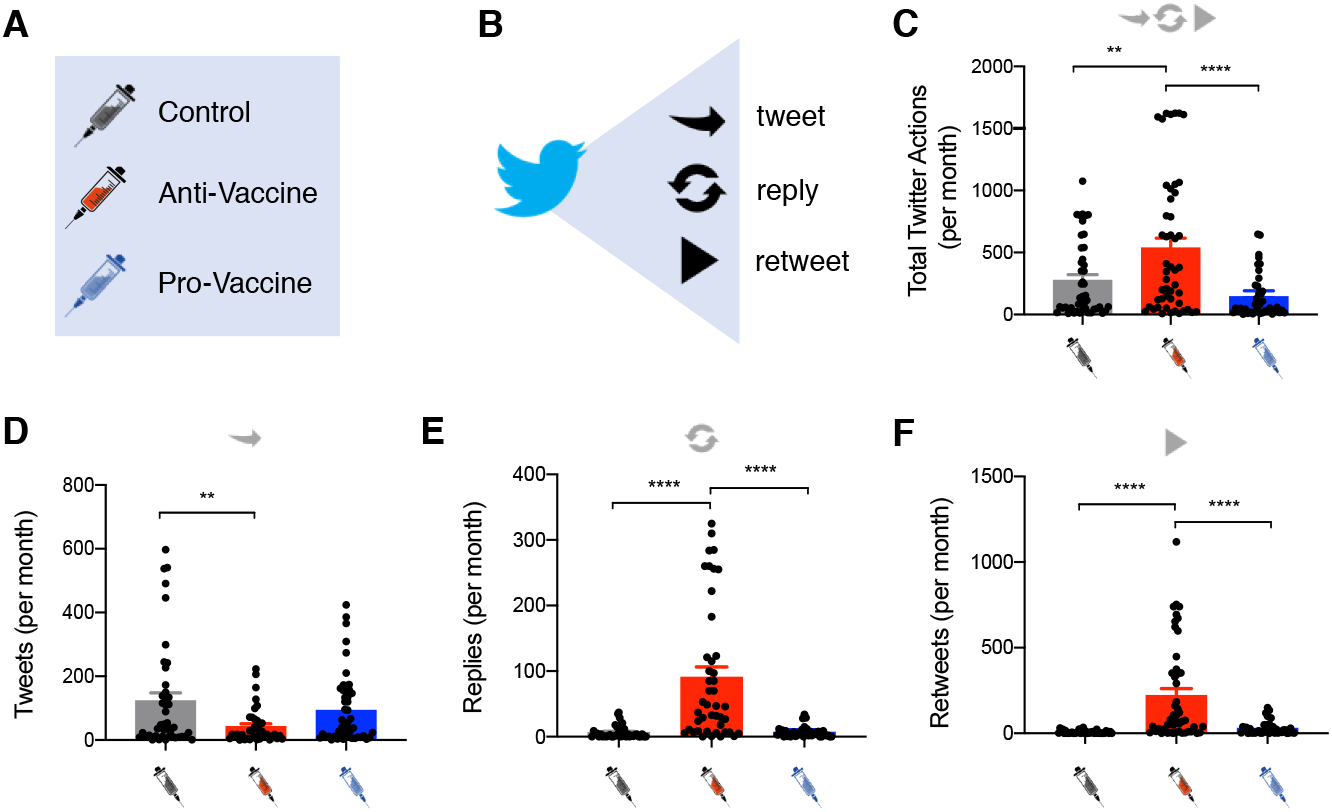
Anti-vaccination supporters are more engaged on Twitter. We analyzed the behavior of three different groups: control (grey), anti-vaccination (red) and pro-vaccination (blue) (**A**). We calculated the number of tweets, replies and retweets per month (**B**). The anti-vaccination group scored the highest number of total Twitter actions (the sum of tweets, replies and retweets) per month (**C**). Anti-vaccination supporters tweeted less than control and pro-vaccination individuals (**D**), but they engaged in more discussion via an increased number of replies (**E**) and Retweets (**F**). Ordinary one-way ANOVA; **p<0.01; ****p<0.0001; Outliers were excluded with ROUT, Q=0.1%; n=50.

## Anti-vaccination support on Twitter is associated with a general belief in conspiracy theories and emotional behaviors

As we have seen, the anti-vaccination community constitutes an echo chamber for misinformed views about vaccines generated by a smaller number of profiles. In order to understand whether these dynamics are established by factors previously associated with vaccine hesitancy *(8, 9, 23)*, we quantified the number of conspiracy theory (CT)-associated contents (tweets and retweets), as well as the number of emotional contents (either depicting emotional situations or adopting emotional language) shared by control, anti-Vaccination and pro-vaccination profiles. Furthermore, we calculated how dedicated the different groups are to share scientific and vaccines-related contents. We found that both pro- and anti-vaccination profiles share a larger number of science- and vaccines-related contents when compared with control profiles (for scientific content: 2.5, 3.4 and 0 per month, respectively; for vaccines-related content: 1.2; 1.5 and 0 per month, respectively) (Fig. 2A, B). Normalization of the aforementioned data for the total number of contents on any given topic indicates that the pro-vaccination group is the most interested in science and vaccines, when compared with anti-vaccination and control groups (Fig. 2A’, B’). Additionally, the anti-vaccination group was the only one circulating conspiracy theories (with an average of 2 contents per month). (Fig. 2C, C’). Most conspiracy theory-related tweets were associated with fake news concerning ruling elites, masonries and techniques of population control – often associated to public figures such as Bill Gates or to ongoing COVID-19 pandemic –, flat earth ideology or pedophilia scandals such as ‘pizzagate’, but also more bizarre ones. The anti-vaccination group shared a larger number of emotional contents per month (and/or content with emotional language) when compared with the pro-vaccination group and control group (1.5, 0.4 and 0 per month, respectively) (Fig. 2D). The normalization of these data for the total number of contents on any given topic shows that anti-vaccination supporters adopt emotional language and/or publish content containing emotional information in 25% of the cases, whereas the pro-vaccination group in only 0.3% of the cases (Fig. 2D’). In line with what was previously reported *(9, 23)*, this suggests that the emotional sphere, which is also connected to the belief in conspiracy theories, is a predominant character of individuals supportive of the anti-vaccination movement. In order to understand whether anti-vaccination contents are associated with conspiracy theories, we calculated the normalized number of vaccines-related contents and correlated it with the number of CT-related contents. As a positive control, we calculated whether the normalized number of science-related contents is correlated with the number of vaccines-related contents published by profiles associated with either the anti- or pro-vaccination groups. As expected, being vaccines-related contents considerable as scientific contents themselves, in both cases there is a clear correlation between the aforementioned factors (R^2^=0.4654; p<0.0001**** and R^2^=0.5924; p<0.0001****, respectively) (Fig. S3). For the anti-vaccine group, there was a strong and significant correlation between the number of published contents against the use of vaccination and the number of published contents concerning conspiracy theories (R^2^=0.7479; p<0.00001****) (Fig. S4A), suggesting that anti-vaccination support can be seen as a part of a bigger problem connected to beliefs in unsubstantiated claims. As pro-vaccine supporters do not share conspiracy theories on Twitter, there is no correlation between these contents and vaccines-related contents (Fig. S4A’). While performing the analysis, we further realized that a large portion of anti-vaccination profiles were sharing contents associated to children, not necessarily in relations to vaccination. For this reason, we decided to quantify the number of children-related content produced in the three groups. In comparison to the control, both anti- and pro-vaccination groups share a higher number of contents associated with children (control: 0; anti-vaccine: 1.2; pro-vaccine: 0.6 contents per month. 0%, 5.7% and 7.3% of the contents concern children, respectively) (Fig. S5). However, we noticed a substantial difference in the communication strategy and topics associated with children in the pro- and anti-vaccination groups. Pro-vaccination supporters generally shared contents depicting happy children after having received a shot whereas anti-vaccination supporters often shared disturbing images of suffering children, or citations of discredited or non-existing physicians about the dangers of vaccines for children. Further, children-related content in this group is also associated with other conspiracy theories about pedophilia scandals, or more generally about sexual and psychological abuses of children.

**Figure 2.**
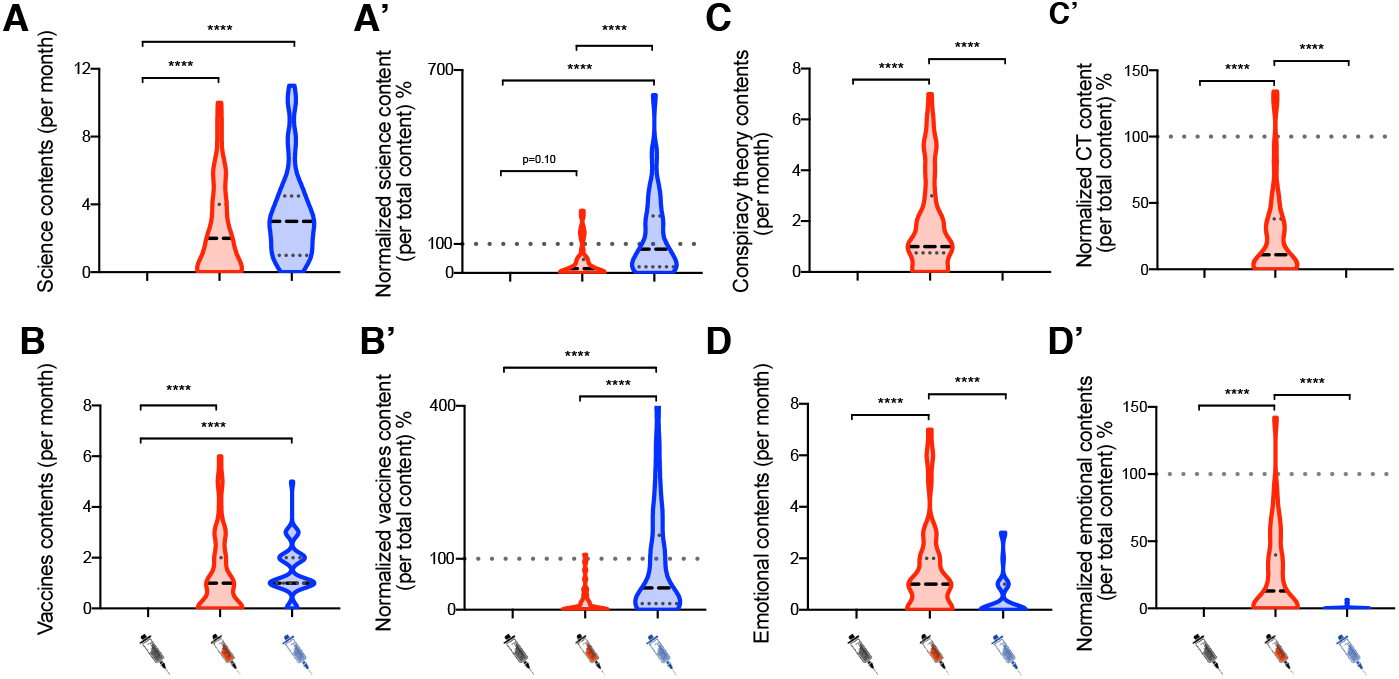
Anti-vaccination supporters are active science and vaccine communicators, share conspiracy theories and emotional content. Both anti- (red) and pro-vaccination profiles (blue) share a larger number of science- and vaccine-related content per month, when compared with control profiles (grey) (**A, B**). We calculated the number of science- and vaccines-related content (tweets and retweets) published in the 24 hours before data analysis and normalized it for the total number of tweets published on average during a single day. 100 percent indicates that all generated contents are estimated to be science- or vaccines-related **(A’, B’)**. Natural fluctuations above 100 percent are due to a variable Twitter activity during the 24 hours prior to data analysis compared to an average day. Anti-vaccination supporters publish conspiracy theories, whereas control and pro-vaccination individuals do not publish this type of material **(C, C’)**. Anti-vaccination supporters share a larger number of tweets and retweets with emotional contents (and with emotional language) compared with the pro-vaccination and control groups (**D, D’**). Ordinary one-way ANOVA; ****p<0.0001; Outliers were excluded with ROUT, Q=0.1%; n=50.

## Emotional language could aid the success of vaccination campaigns

As we previously described, anti-vaccination supporters share emotional contents with the use of emotional language. In order to understand whether this language is necessary for the success of the movement, we decided to perform an analysis of the most used words by the three different groups. We considered the 5 most used words for each individual profile and calculated the most used words for each individual group. Following normalization against the words predominantly used by control profiles, we identified a list of 10 words strongly associated with anti- and pro-vaccination groups (Fig. 3A, A’). As expected, the word “vaccine(s)” is the most represented in both groups, confirming that our initial criteria for inclusion were reasonable. To further highlight the differences between the two groups, we normalized the most used words in the two groups against each other (Fig. 3B). Here we found that the most relevant words in the anti-Vaccination group are “President”, “God”, “People”, and “Masks”. In contrast, pro-vaccination profiles preferentially included words such as “Help”, “Health”, “Thanks” or “Research”. In order to better determine the interests of the different groups, we clustered words according to topics, and found that anti-vaccination profiles are the most engaged in political discussion, with nearly a 6-fold increase compared with the pro-vaccination group (Fig. 3C). Finally, we analyzed whether the use of emotional contents and language is associated with increased engagement, measured as the sum of likes, replies and retweets on each individual tweet, but found no significant correlation between the two factors for the anti-vaccination group (Fig. 3D). On the contrary, the pro-vaccine group showed a significant correlation between the two aforementioned factors (Fig. 3D’), suggesting that the use of emotional language could aid the success of the pro-vaccination communication strategy online.

**Figure 3.**
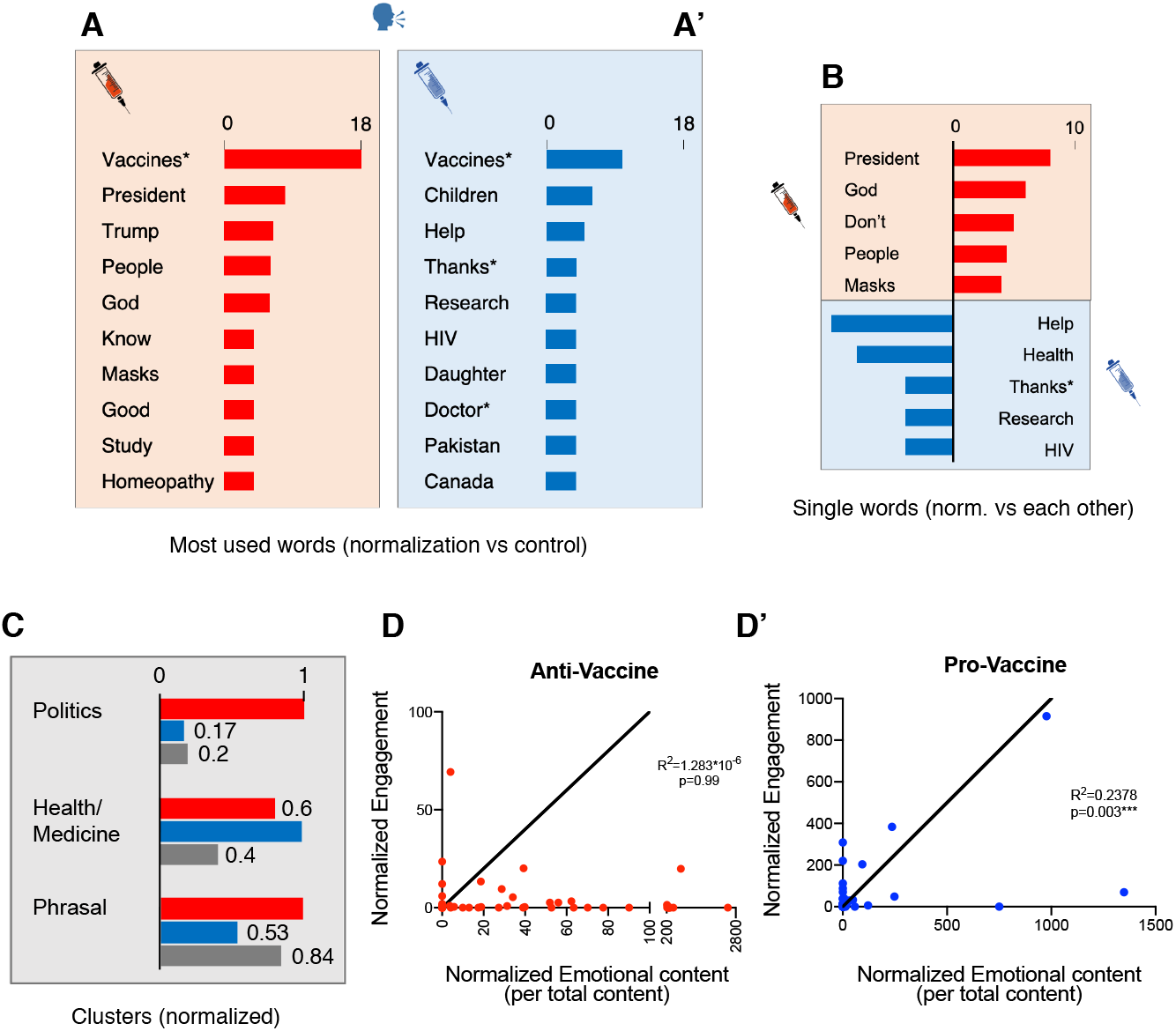
The anti-vaccination group utilizes emotional language, but this does not determine the success of their tweets (engagement). Most used words on Twitter by the anti-(red) and pro-vaccination groups (blue) normalized against the words predominantly used by the control-group (grey). Asterisks* indicate that words have been clustered (e.g. “vaccine” and “vaccines” are scored as a single word). n(profiles analyzed)=42. Max=18 indicates that particular word is used 18-times more in that specific group, when compared with the control. (**A, A’**). Most used words by anti- and pro-vaccination profiles normalized against each other. Asterisks* indicate clustered words. n(profiles analyzed)=42 (**B**). Words are clustered for topic and normalized, with the value of 1 being assigned to the group utilizing the cluster of words the most. The most relevant clusters are shown. Words related to politics are greatly enriched in the anti-vaccination group; words related to health and medicine are predominantly used by pro- and anti-vaccination profiles, when compared with the control; phrasal words are underrepresented in the pro-vaccination group. Asterisks* indicate clustered words. (**C**). For the anti-vaccination group, the normalized number of emotional contents (relative to the total number of contents generated by a given profile) does not correlate with the number of engagements received on average for a single tweet (R^2^=1.293*10^−6^; p=0.99); n=50 (**D**). Conversely, pro-vaccination profiles tweeting emotional content produce more engaging contents (R^2^=0.2378; p=0.003); n=50 (**E**).

## Pro-vaccination supporters are more interested in their own education and profession

Previous studies showed that education might increase confidence in vaccine importance and effectiveness *(32)*. However, different studies reached different conclusions on whether education plays a role in shaping vaccination choice *(33, 34)*. We therefore decided to quantify the number of profiles associated with the three groups that declared their education or profession status. This analysis does not determine whether education plays a factor in shaping vaccination choice. However, it determines whether holding a position in the vaccination debate is associated with a self-perceived relevance of education. To determine whether the source of information is of relevance in this context, we scored the number of profiles publicly declaring their name and surname, together with a seemingly real profile picture. Here we show that the great majority of pro-vaccine profiles declares their identity when compared with the control (64% vs 30%, respectively), and that anti-vaccination supporters are particularly reluctant to do so (only 16%) (Fig. S6A). Similarly, education and/or profession in the Twitter headline was declared 32% of the times in the pro-vaccination group, compared with 10% and 6% in the control and anti-vaccination group, respectively (Fig. S6B).

## The pro-vaccination group produces the most engaging contents

As we have discussed so far, the success of the anti-vaccination message is not determined by a larger production of original contents, and the use of emotional language is a structural component of this group that does not influence engagement. Here we show that the pro-vaccination group produces the most engaging contents, whereas the anti-vaccination group produces the least engaging contents (Pro-vaccine: 15.2 engagement per tweet; control: 3.7; anti-vaccine: 0.8) (Fig. 4A), and the average engagement per tweet is 19.9 times higher than for the anti-vaccination group (and 5.5 times higher than for the control) (Fig. 4B). On average, pro-vaccination profiles are also those with a larger number of followers, when compared with control and anti-vaccination groups (mean: 1841; 605 and 338 followers, respectively) (Fig. 4C). Here we show that contents published by the pro-vaccination group are more engaging than contents produced by the majority of anti-vaccination profiles. In light of this results, we hypothesized that the success of the anti-vaccination movement is likely driven by a stronger sense of community, built around common interests (besides vaccines), and based on personal perspective and emotional language. We therefore hypothesized the existence, in this community, of a pull of influencers producing the most engaging contents, with the vast majority of anti-vaccination profiles functioning as the recipient and echo chamber for these messages, whereas novel contents produced by these profiles receive little attention compared with contents generated by an average pro-vaccination supporter (illustrative scheme in Fig. 4D).

**Figure 4.**
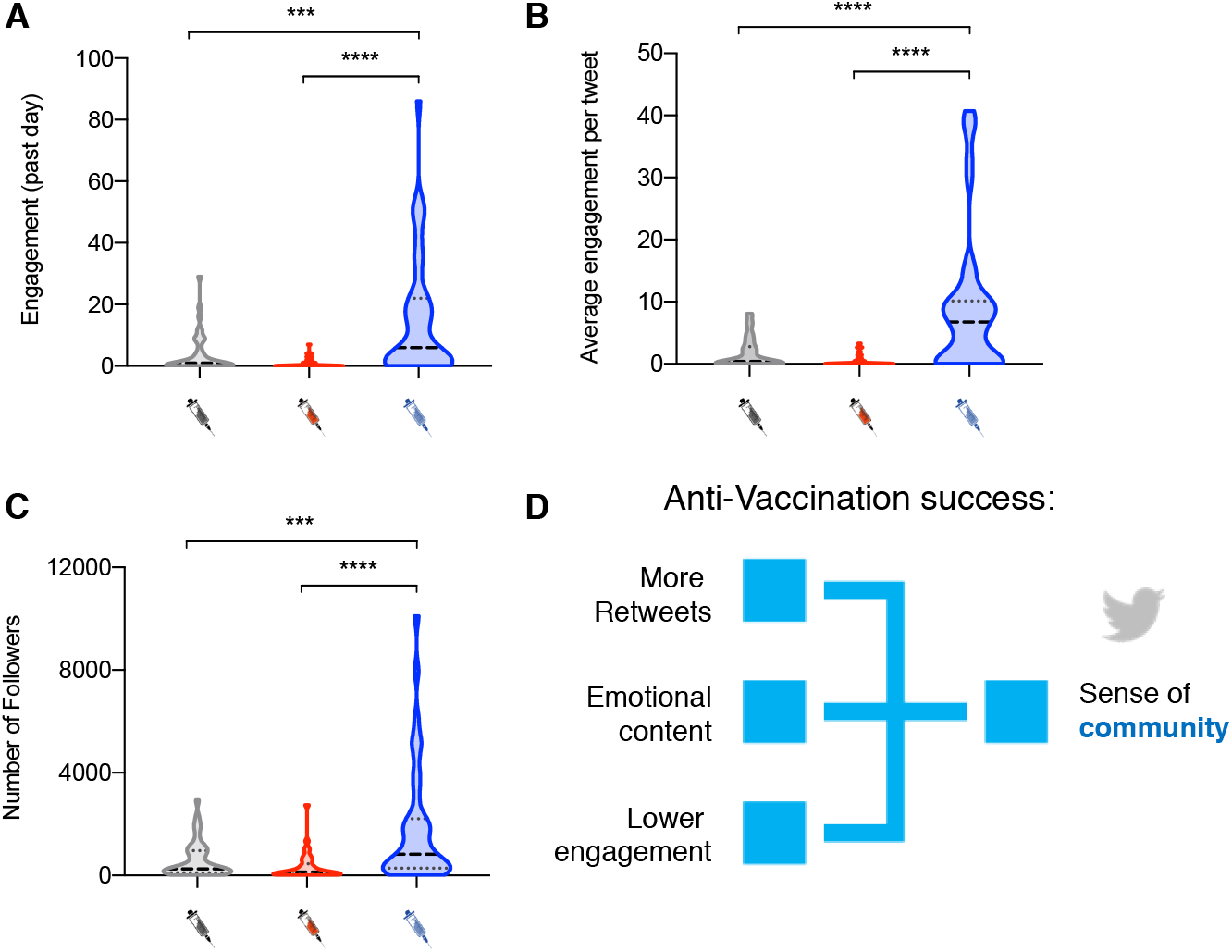
Pro-vaccination profiles have more followers and produce more engaging content. Pro-vaccination profiles (blue) generate more engagement in one day when compared with the control (grey) and anti-vaccination groups (red) (**A**), and normalization shows they produce more engaging content irrespectively of the number of contents generated in a given day (**B**). Pro-vaccination profiles have a larger number of followers when compared with the control and anti-vaccination groups (**C**). Hypothetical model to illustrate the results described so far. Anti-vaccination supporters are more engaged on Twitter, as they retweet contents more often than control and pro-vaccination profiles. They also share emotional content, although they generally produce less engaging content than their pro-vaccination counterparts. Despite the use of emotions as a tool to convey their message, given the lower engagement of anti-vaccination tweets, we hypothesized that a sense of community driven by common interest is key for the success of the anti-vaccination movement online (**D**). Ordinary one-way ANOVA; ***p<0.001; ****p<0.0001; Outliers were excluded with ROUT, Q=0.1%; n=50.

## Anti-vaccination supporters are engaged in a virtual community led by Donald Trump and other influencers

In order to determine whether the success of the anti-vaccination movement is due to the existence of a community of engaged individuals driven by a pull of influencers with large follows, we retrieved, for each individual profile of both the anti- and pro-vaccination group (n=42 each), the 10 most retweeted profiles, and included them in our analysis. We scored the number of connections (edges; E) they established with each other by building a Twitter web with Cytoscape *(35)*. The pro-vaccination (Fig. 5A) and anti-Vaccination Twitter webs (Fig. 5B), scaled 1:1, show the extent of the ramifications of the latter in comparison to the former (Fig. 5A, B). The size of each node (profile) is scaled linearly depending on the number of edges. Color is also indicative of the number of edges, and thus of the relevance of the node in the web (no color: E<2; yellow: 2≤E≤4; orange: 5≤E ≤9; red: E ≥10). Close ups (not equally scaled, for better readability) show the most relevant section of the pro- and anti-vaccination webs (Fig. 5A’, B’). The average number of neighbors in the web is 1.45-folds higher in the anti-vaccination web when compared with the pro-vaccination web (2.8 and 2 neighbors, respectively), the clustering coefficient is also higher in the anti-vaccination web (0.021 and 0.007, respectively), as well as the density of the network (0.005 vs 0.003) and the characteristic path length (1.6 vs 1.4) (Fig. 5C). In addition, the pro-vaccination web has a similar number of nodes and edges, whereas the anti-vaccination web has a larger number of edges than nodes. Therefore, the number of edges per nodes, which indicates the number of existing connections for each individual profile in the web, is much larger in the anti-vaccination group when compared with the pro-vaccination group (1.51 vs 1.02 connections per profile, respectively) (Fig. S7), confirming that anti-vaccination supporters are well-connected in a community. Furthermore, with an E≥5 cut-off, we identify only one large influencer in the pro-vaccination web (the World Health Organization, E=5), whereas, according to the same criterium, we identify 14 large influencers, with the largest one being US President Donald Trump (E=26), 5.2 times more relevant than the World Health Organization in the pro-vaccine web. Other influencers include Trump’s family members, politicians and public figures known to support his presidency, as well as individuals and unverified popular profiles that are fully committed to the vaccination issue. Therefore, here we identify the pull of relevant influencers that are likely to determine the opinion about vaccine of a large number of people. These influencers include Trump – who is himself a proven anti-vaccination supporter, and others, such as activist Charlie Kirk or vaccine-denier Eileen Iorio.

**Figure 5.**
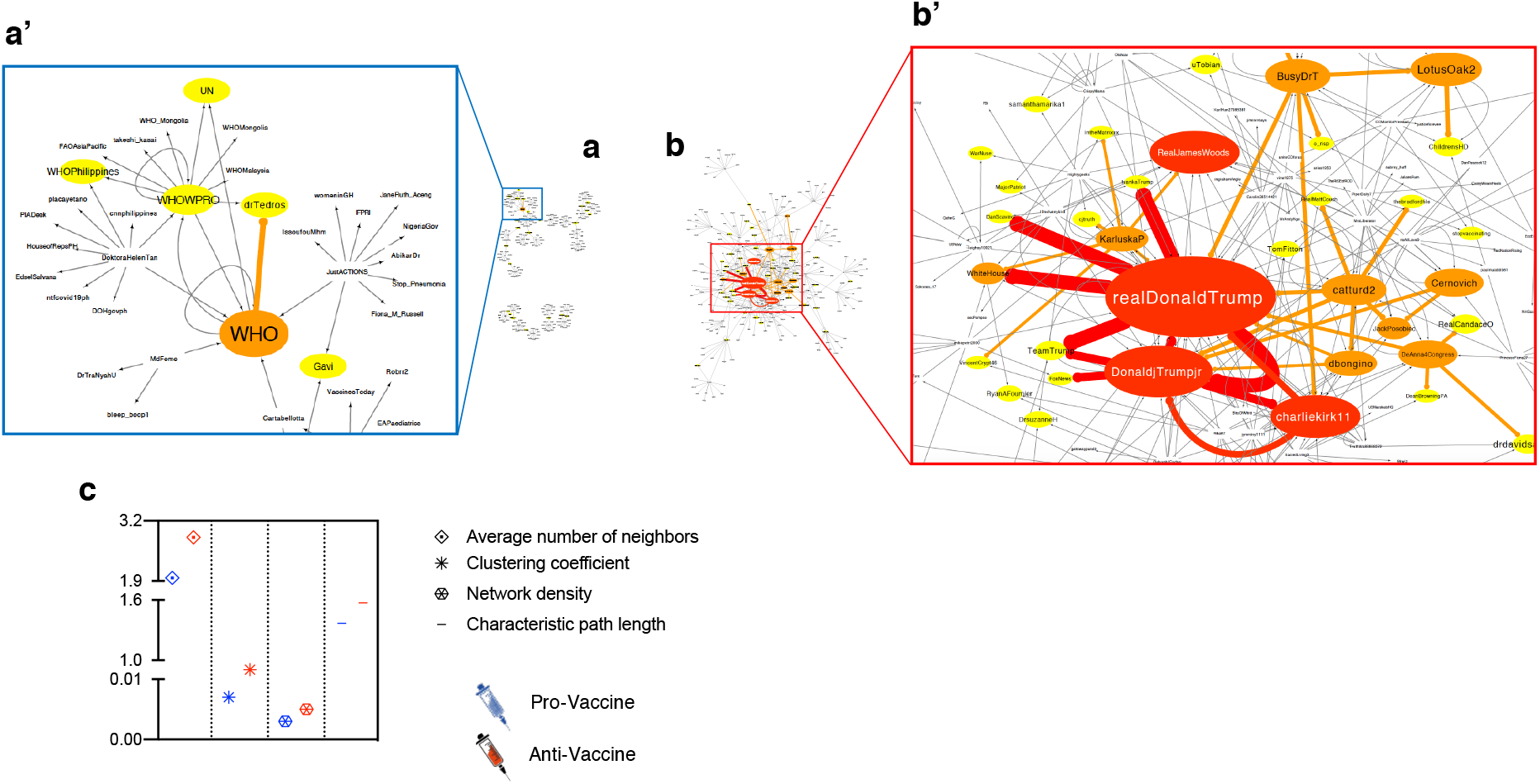
Anti-vaccination profiles establish a well-connected community sharing contents produced by a pull of influencers, whose most prominent exponent is Donald Trump. The pro-vaccination Twitter web (**A**). Close up of the most relevant portion of the pro-vaccination web, which highlights the World Health Organization as the main influencer for the pro-vaccination group (**A’**). The anti-vaccination Twitter web (**B**). Close up of the most relevant portion of the anti-vaccination web, which highlights Donald Trump, its political entourage and public figures supporting his presidency as the main influencers for the anti-vaccination group (**B’**). The pro-vaccination and anti-vaccination Twitter webs are scaled 1:1 (**A, B**). For better readability, close up representations of the pro- and anti-vaccination webs are not equally scaled. Yellow color represents Twitter profiles (nodes) with 2 to 4 anti-vaccination profiles preferentially retweeting their contents within the top 10 most retweeted users (edges; 2≤ E ≤4; n=42). Orange nodes represent profiles with 5 to 9 edges (5≤ E ≤9; n=42), whereas red nodes indicate profiles with more than 10 connecting edges (E ≥10; n=42). Size of the nodes is linearly scaled depending on the number of edges connecting the node (**A-B’**). The average number of neighbors in the web, the clustering coefficient, the density of the network and the characteristic path length of the anti-vaccination (red) web is greater than the pro-vaccination counterpart (blue) (**C**). Graphical representation and web parameters were generated with Cytoscape. Graphical representation of the main influencers in the pro- and anti-vaccination Twitter webs (threshold: E ≥5; n=42). The size of the profile picture and the name assigned to the Twitter profile are linearly scaled for the number of edges.

## The Anti-vaccination community and political implications

In this paper we show that anti-vaccination supporters produce fewer original contents on Twitter but share more contents than users belonging to the pro-vaccination or control group. However, we also show that the average engagement, calculated as the sum of comments, likes and retweets received by an anti-vaccination tweet, is extremely low when compared with tweets published by pro-vaccination profiles. This indicates that the majority of anti-vaccination supporters is unlikely to influence vaccination choice for a large number of individuals. Instead, we show that the success of the anti-vaccination movement online is likely based on common beliefs and interests, through which users establish a well-connected community and constitute an echo chamber for contents generated by a smaller fraction of profiles. We define these latter users as anti-vaccination influencers. We identify US President Donald Trump as the main influencer in the anti-vaccination web. Despite him not having published direct anti-vaccination tweets in recent times, Donald Trump consistently shared anti-vaccine contents in the past, often associating vaccines to autism. In addition, his political position, and personal beliefs, remain strongly related to anti-vaccination positions. Besides Trump, we identify his son Donald Trump Jr, Charlie Kirk, a popular evangelical Christian and Republican activist who supports Trump’s presidency, James Wood, a popular actor and producer who is also a strong supporter of Trump – to be among the largest influencers in the anti-vaccination network. Among others, there are also profiles fully dedicated to spread the anti-vaccination message online, including authors of books on the dangers of vaccines, and non-verified profiles including Catturd2, a ‘cat’ who defines itself as “The MAGA turd who talks shit”. Interestingly, in a recent study Trump was identified as the largest driver of the COVID-19 infodemic *(26)*, underlining the necessity of a scientific movement that prompts politicians to base their campaigns on evidence-driven policies.

## The polarization of the anti-vaccine debate

Here we demonstrate that anti-vaccination supporters share conspiracy theories, and that anti-vaccine messages can be for a substantial part be considered as conspiracy theories themselves. This process is likely driven by the polarization of social media feed, where users are exposed to information, news and views identified by algorithms as close to their interests. Conspiracy theories of various kinds, as well as anti-vaccination beliefs and political extremism tend to be associated with each other *(36, 37)*. As we previously mentioned, Donald Trump, despite being an anti-vaccination supporter, has not discussed vaccination issues in similar terms during his presidency. Nonetheless, he retains the indirect ability to influence the great majority of individuals associated with the anti-vaccination movement. Simply put, due to the polarization of the debate on social media, sharing or reading conservative tweets could increase the chance that a hesitant person gets in touch with anti-vaccination beliefs. In line with this, it was previously shown that anti-vaccine users form a polarized network with little to no interaction with outsiders, in which users strengthen their positions by sharing each other’s contents *(38-40)*. We therefore strongly encourage social media to change the polarized way they present information to users to halt the anti-vaccination infodemic and increase debate between communities. In addition, social media could target these influencers in different ways. These could include ‘shadow bans’ for science-based contents – which could force a tweet’s organic reach to drop (i.e. a small number of people would read the content); info banners for tweets containing unverified information about medically-sensitive topic could also be effective tools to limit the spread of misinformation about vaccines. Finally, we encourage social media and the scientific community to discuss the possible introduction of science knowledge tests, which could be required for users that intend to share contents containing medically-sensitive information. This would restrict a user’s ability to tweet and share misinformed views about vaccines, without imposing an a priori restriction of individual freedom of speech. Furthermore, as the strength of the anti-vaccination movement relies on the structure of its community, health organizations should consider lobbying indirect anti-vaccination influencers to become active pro-vaccination communicators.

## Towards a better communication strategy for vaccinations

Finally, here we show that the use of people-centered, first-person narratives with emotional language could aid the communication strategy of pro-vaccine health organizations and individuals. The power of first-person narratives over population-based statistical evidence could be due to an effect known in psychology as “psychic numbing”, according to which the higher the number of people involved in a disaster and the least people feel empathic about it. Personal stories, involving first person narratives, are more attractive and stimulate empathic responses more efficiently *(41-43)*. Given that this type of communication seems to be a structural component within the anti-vaccination community, it may be required for users to build strong connections. We therefore encourage health organizations to adopt a less sterile, technical language when communicating with the general public. This language should be scientifically sound, but also simple, emotional and understandable. At the same time, adopting a pro-active long-term strategy for increasing the general public’s science literacy and ability to read and understand basic scientific information will be an important complementary strategy.

## Supporting information

Anti-Vaccination network

Pro-vaccination network

## Data Availability

All data are available in the manuscript.

## Acknowledgments

We thank Anna Stelling for proofreading the manuscript. Funding: This work was supported by the Swiss National Science Foundation. Authors contributions: FG and NBA ideated this research project. FG carried out experimental work. FG and NBA wrote the manuscript. The authors declare no conflicts of interest. All data is available in the manuscript or in the supplementary materials.

## Statement

the authors declare no conflicts of interest.

## Contribution

F.G. and N.B. conceived the idea of the project. F.G. performed the research analysis. F.G. and N.B. wrote the manuscript.

## References

1. S. Tafuri, M.S. Gallone, M.G. Cappelli, D. Martinelli, R. Prato, C. Germinario. Addressing the anti-vaccination movement and the role of HCWs. Vaccine. Aug 27;32(38):4860-5 (2014).

2. J. Hammond. Vaccine confidence, coverage, and hesitancy worldwide: A literature analysis of vaccine hesitancy and potential causes worldwide. Senior theses. 344 (2020).

3. F. DeStefano. Vaccines and autism: evidence does not support a causal association. Clinical Pharmacology & Therapeutics 82(6): 756–9 (2007).

4. M.F. Johnson, N. Velasquez, N.J. Restrepo, R. Leahy, N. Gabriel, S. El Oud, M. Zheng, P. Manrique, S. Wutchy, Y. Lupu. The online competition between pro- and anti-vaccination views. Nature. 582, 230–233 (2020).

5. T. Burki. The online anti-vaccine movement in the age of COVID-19. The Lancet Digital Health 2,10,504–505 (2020).

6. A.B. Amin, R.A. Bednarczyk, C.E. Ray, K.J. Melchiori, J. Graham, J.R. Huntsinger, S.B. Omer. Association of moral values with vaccine hesitancy. Nature Human Behaviour 1, 873–880 (2017).

7. L. Prior. Belief, knowledge and expertise: the emergence of the lay expert in medical sociology. Sociology of Health & Illness 25; 44–57 (2003).

8. A. Kata. A postmodern Pandora’s box: Anti-vaccination misinformation on the Internet. Vaccine 28; 1709–1716 (2010).

9. Z, Xu, L. Ellis, L.R. Umphrey. The easier the better? Comparing the readability and engagement of online pro- and anti-vaccination articles. Health Education and Behavior 46,5; 790–797 (2019)

10. M.U. Hornsey, E.A. Harris, K.S. Fielding. The psychological roots of anti-vaccination attitudes: A 24-nation investigation, Health Psychology 37(4), 307–315 (2018).

11. A. Tsuda, K.R. Muis. The anti-vaccination debate: a cross-cultural exploration of emotions and epistemic cognition. Advances in Social Sciences Research Journal 5(9) (2018).

12. C. Betsch, P. Schmid, D. Heinemeier, L. Korn, C. Holtmann, R. Böhm. Beyond confidence: Development of a measure assessing the 5C psychological antecedents of vaccination, PLoS ONE 13(12) (2018).

13. T. Mitra, S. Counts, J.W. Pennebaker. Understanding anti-vaccination attitudes in social media. Proceedings of the Tenth International AAAI Conference on Web and Social Media (2016).

14. A.L. Schmidt, F. Zollo, A. Scala, C. Betsch, W. Quattrociocchi. Polarization of the vaccination debate on Facebook. Vaccine 36; 3606–3612 (2018).

15. A. Hussain, S. Ali, M. Ahmed, S. Hussain. The anti-vaccination movement: a regression in modern medicine, Cureus 10(7) (2018).

16. B.L. Hoffman, E.M. Felter, K. Chu, A. Shensa, C. Hermann, T. Wolynn, D. Williams, B.A. Primack. It’s not all about autism: The emerging landscape of anti-vaccination sentiment on Facebook. Vaccine 37,16 2216–2223 (2019).

17. N. Smith, T. Graham. Mapping the anti-vaccination movement on Facebook. Information, Communication & Society 22; 1310–1327 (2017).

18. A. Kata. Anti-vaccine activists, Web 2.0, and the postmodern paradigm – An overview of tactics and tropes used online by the anti-vaccination movement. Vaccine 30,25 3778–3789 (2012).

19. L. Chou, C. Tucker. Fake News and advertising on social media: a study of the anti-vaccination movement. National Bureau of Economic Research 25223 (2018).

20. A. Evrony, A. Caplan. The overlooked dangers of anti-vaccination groups’ social media presence. Human Vaccines & Immunotherapeutics. 13,6; 1475–1476 (2017).

21. D.J. Opel, J.A. Taylor, R. Mangione-Smith, C. Solomon, C. Zhao, S. Catz, D. Martin. Validity and reliability of a survey to identify vaccine-hesitant parents. Vaccine 29(38): 6598–605 (2011).

22. K. Madden, X. Nan, R. Briones, L. Waks. Sorting through search results: a content analysis of HPV vaccine information online. Vaccine 28;30(25): 3741–6. (2012).

23. C. Betsch, F. Renkewitz, T. Betsch, C. Ulshöfer. The influence of vaccine-critical websites on perceiving vaccination risks. Journal of Health Psychology 15,3; 446–455 (2010).

24. C. Betsch, F. Renkewitz, N. Haase. Effect of narrative reports about vaccine adverse events and bias-awareness disclaimers on vaccine decisions: a simulation of an online patient social network. Med Decis Making 33(1):14-25 (2013).

25. A. Allam, P.J. Schulz, K. Nakamoto. The impact of search engine selection and sorting criteria on vaccination beliefs and attitudes: two experiments manipulating Google output. J Med Internet Res 16(4):e100(2014).

26. S. Evanega, M. Lynas, J. Adams, K. Smolenyak. Quantifying sources and themes in the COVID-19 ‘infodemic’. Cornell Alliance For Science (2020).

27. H. Alcott, M. Gentzkow. Social media and fake news in the 2016 election. Journal of Economic Perspectives 31(2): 211–36. (2017).

28. Y. Gorodnichenko, T. Pham, O. Talavera. Social media, sentiment and public opinions: evidence from #Brexit and #USElection. National Bureau of Economic Research (2018).

29. M. Hänska, S. Bauchowitz. Tweeting for Brexit: how social media influenced the referendum. Published in J. Mair, T. Clark, N. Flower, R. Snoody, R. Tait. Brexit, Trump and the media. Abramis Academic Publishing, 31–35 (2017).

30. J. Kennedy. Populist politics and vaccine hesitancy in Western Europe: an analysis of national-level data. European Journal of Public Health, 29(3): 512–516 (2019).

31. The COCONEL Group. A future vaccination campaign against COVID-19 at risk of vaccine hesitancy and politicisation. The Lancet Infectious Diseases. 20(7): 769–770 (2020).

32. H.J. Larson, A. de Figueiredo, Z. Xiahong, W.S. Schulz, P. Verger, I.G. Johnston, A.R. Cook, N.S. Jones. The state of vaccine confidence 2016: Global insights through a 67-country survey. EBioMedicine 12:295–301 (2016).

33. K.F. Brown, J.S. Kroll, M.J. Hudson, M. Ramsay, J. Green, S.J. Long, C.A. Vincent, G. Fraser, N. Sevdalis. Factors underlying parental decisions about combination childhood vaccinations including MMR: a systematic review. Vaccine 28(26):4235–48 (2010).

34. H.J. Larson, C. Jarrett, E. Eckersberger, D.M.D. Smith, P. Paterson. Understanding vaccine hesitancy around vaccines and vaccination from a global perspective: a systematic review of published literature, 2007-2012. Vaccine 32(19):2150–9 (2014).

35. P. Shannon, A. Markiel, O. Ozier, N.S. Baliga, J.T. Wang, D. Ramage, N. Amin, B. Schwikowski, T. Ideker. Cytoscape: a software environment for integrated models of biomolecular interaction networks. Genome Res 13(11): 2498–504 (2003).

36. M. J. Wood, K.M. Douglas, R.M. Sutton. Dead and alive: Beliefs in contradictory conspiracy theories. Social Psychological and Personality Science 3(6): 767–773 (2012).

37. J. van Prooijen, A.P.M. Krouwel, T.V. Pollet. Political extremism predicts belief in conspiracy theories. Social Psychological and Personality Science 6(5): 570–578 (2015).

38. E. Milani, E. Weitkamp, P. Webb. The visual vaccine debate on Twitter: A social network analysis. Media and Communication 8(2): 364–375 (2020).

39. G. Bello-Orgaz, J. Hernandez-Castro, D. Camacho. Detecting discussion communities on vaccination in twitter. Future Generation Computer Systems 66: 125–136 (2017).

40. X. Yuan, A.T. Crooks. Examining online vaccination discussion and communities in Twitter. SMSociety ‘18: Proceedings of the 9th International Conference on Social Media and Society 197–206 (2018).

41. P. Slovic, D. Västfjäll, A. Erlandsson, R. Gregory. Iconic photographs and the ebb and flow of empathic response to humanitarian disasters. Proc Natl Acad Sci U S A 114(4):640-644 (2017).

42. D. Västfjäll, P. Slovic, M. Mayorga, E. Peters. Compassion fade: affect and charity are greatest for a single child in need. PLoS One 9(6):e100115 (2014).

43. S.R. Maier, P. Slovic, M. Mayorga. Reader reaction to news of mass suffering: Assessing the influence of story form and emotional response. Journalism 18(8):1011-1029 (2017).

