## Supplementary Material and Figures for "The anti-vaccination infodemic on social media: a behavioral analysis"

### Materials and Methods

#### Definition of pro-, anti-vaccination and control users

Profiles belonging to the pro-, anti-vaccination or control group were initially automatically identified for their use of hashtags associated with the respective groups, and then manually screened to ensure users truly expressed opinions in line with expectations for any given group. Control individuals were identified for their use of #control hashtags, which were selected via an online Random Word Generator tool (available at https://randomwordgenerator.com). Each control profile was selected via a different randomly generated word. pro-vaccination individuals were identified for their use of the #vaccineswork hashtag, whereas anti-vaccination profiles were identified for their use of either the #vaccineskill or the #vaccinesharm hashtag.

#### Scoring the number of tweets, replies and retweets

We manually calculated the number of tweets, replies and retweets published in the previous 24 hours for all the 50 profiles we analyzed in each group. This included the number of science-, vaccines-, conspiracy theory- and children-related tweets, as well as ‘emotional’ tweets. In order to determine the overall number of tweets, replies and retweets, we used the freely available tool online TweetStats (www.tweetstats.com). After feeding a Twitter username, the software returns the number of contents generated on average in a month since the profile was initially set up, as well as the percentage of replies and retweets. In order to calculate the normalized percentage of tweets concerning a given topic against the overall number of tweets, we divided the number of tweets concerning a topic of interest – which were generated in the 24 hours before the analysis – for the number of average tweets per day. This number generally fluctuates between 0% (no contents of the analyzed topic) and 100% (all contents are associated to a given topic). However, this percentage can occasionally exceed 100% due to fluctuations between the number of tweets published on average in a day and the actual number of tweets published in the 24 hours prior to analysis.

### Statistical analysis

Ordinary one-way ANOVA was used to compare the number of contents, tweets, replies and retweets between the different groups, as well as the differences between the number of retweets per tweet, the number of conspiracy theories-associated contents, the number of emotional tweets or children-related tweets, the total engagement per day, the average engagement per tweet and the number of followers. The statistical analysis was preceded by the elimination of behavioral outliers (excluded with ROUT, Q=0.1%). Behavioral outliers predominantly included profiles sharing a vast number of contents per day. The Chi-square test was used to determine differences in users' behavioral patterns and in particular to determine whether users belonging to different groups would be more or less prone to disclose personal information (name, surname and personal picture), their education or profession status. In general, 50 profiles were analyzed for each individual group and experimental analysis, unless differently specified.

### Language analysis

Using TweetStats, we retrieved the 5 most used words on Twitter for each individual profile belonging to each group (n=42) and compiled a list of the most used words for each group. We assigned a value to each word, depending on how often it was observed to be among the top 5 words used by a profile. For instance, a score of 42 indicates that 100% of analyzed profiles included the word of interest among the 5 most used words on Twitter, whereas 0 indicates that none of the profiles used that particular word often enough. As most of the words were not unique, under- or over-represented in each group, we performed two normalization analyses. The first compared the most used words in the pro- and anti-vaccination groups with words predominantly used by control profiles, and the second compared the most used words in the pro- versus anti-vaccination group. For example, a value of 18 indicates that a particular word has been used 18-times more in one group when compared with another one (such as the word "vaccines in the anti-vaccination group, when compared with the control). If a word was not used in a given group, we arbitrarily doubled the number of times the word was used in the comparison group. In the aforementioned analyses, words were clustered for plurals or variations of the same word with identical meaning. For example, the word "vaccine" and "vaccines" were considered as one variable, as well as the words "Dr", "Doctor" and "Doctors". Words were further clustered per topic. For instance, the category "politics" included words such as "Trump", "Democrats", "Conservative" or "elections"; similarly, the cluster "phrasal" included "Don't", "I'm", "We", etc. As before, we determined whether this clusters are over- or under-represented among different study groups by performing a comparison between the most used clusters in one group versus another group.

### Engagement

Engagement was calculated as the sum of likes, comments and retweets. The average engagement was determined dividing the total engagement for the total amount of tweets published in a given time.

### Network generation and analysis

In order to generate pro- and anti-vaccination networks, we considered n=42 profiles for our analysis. For each of these profiles, assigned to one of the two groups, we used TweetStats to retrieve the profiles of the 10 most retweeted users. This allowed us to identify a larger number of individuals, directly or indirectly involved in the anti-vaccination community. Our analysis considered profiles regardless of their personal position in the vaccination debate, assuming that overrepresented profiles in the network would be associated to the anti-vaccination or pro-vaccination community, respectively. We generated the networks with Cytoscape and retrieved the average number of neighbors, the clustering coefficient, the density of the network and the characteristic path length from the Cytoscape Analyzer Tool. In order to optimize the graphical representation of the webs, we removed clusters of individuals that were not connected with other clusters. We further highlighted in yellow those profiles with a number of edges (connections with other profiles) between 2 and 4, in orange those with a number of edges between 5 and 9, and in red those with 10 or more edges. The size of the node (profile) in the web was also linearly scaled depending on the number of connections. Finally, in order to determine the content-based connections among influencers and between influencers and the community at large, we analyzed the 10 most retweeted profiles for each profile with more than 5 edges and included them in our analysis.

### Education and personal information

In order to define whether a profile was trackable, and ascribable to a real person, we scored the number of individuals publicly declaring their name, surname and utilizing a profile picture of a seemingly real person (n=42 for each group). We defined a profile as trackable when all these criteria were met, and not trackable when at least one of the above-mentioned criteria was not met. “Not defined” (nd) was assigned when the judgment could not be made, for instance when profiles represented institutions without a verification badge. Further, we scored whether profiles indicated either their education or profession status in their Twitter headline. “Yes” indicates that the profile declares either her education or profession publicly, whereas “No” indicates that neither of the two is indicated, and “nd” is assigned when the judgement could not be made. For instance, when profiles represented institutions without a verification badge.

### Supplementary Figures

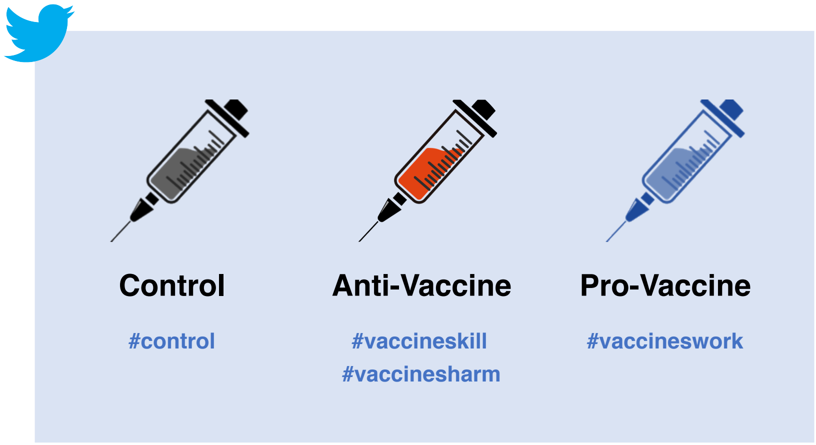


**Supplementary Figure 1. Definition of the study groups and relative hashtags.** We classified profiles in three groups: control (grey), anti-Vaccine (red) and pro-Vaccine (blue). Profiles (n=50 for each group) were identifying automatically through the use of hashtags. Control profiles were selected for their use of randomly selected hashtags, anti-vaccination profiles for their use of widely chosen hashtags in the community (#vaccineskill and #vaccinesharm), whereas Pro-vaccine profiles were selected for their use of the #vaccineswork hashtag.

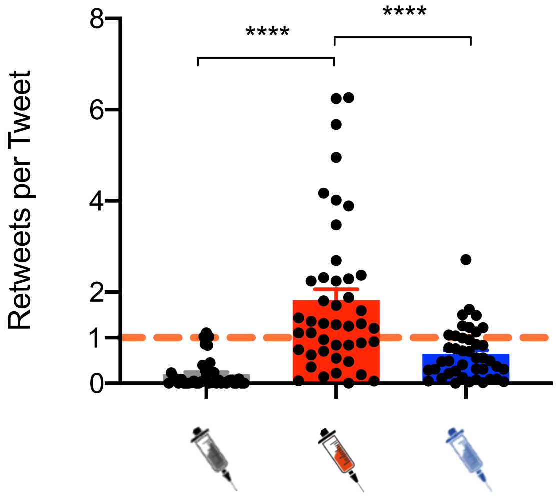


**Supplementary Figure 2. Anti-Vaccination profiles retweet more than they tweet.** Profiles belonging to the control and pro-vaccination groups tweet more than they retweet (1 indicates an equal number of retweets per tweet on average in a month), whereas anti-vaccination profiles retweet more than they tweet. Ordinary one-way ANOVA; ****p<0.0001; Outliers were excluded with ROUT, Q=0.1%; n=50.

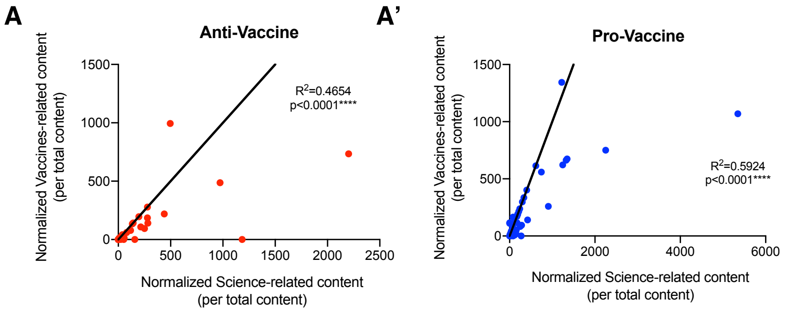


**Supplementary Figure 3. The number of vaccines- and science-related contents shared by anti- and pro-vaccination profiles are correlated.** For both the anti-vaccination group (red) (**A**) and the pro-vaccination group (blue) (**A'**), the higher the normalized number of science-related contents generated or shared (for the overall number of contents generated on any given topic), the larger the number of normalized vaccines-related tweets and retweets ($R^2$=0.464 and $R^2$=0.5924 respectively; ****p<0.0001; Outliers were excluded with ROUT, Q=0.1%; n=50).

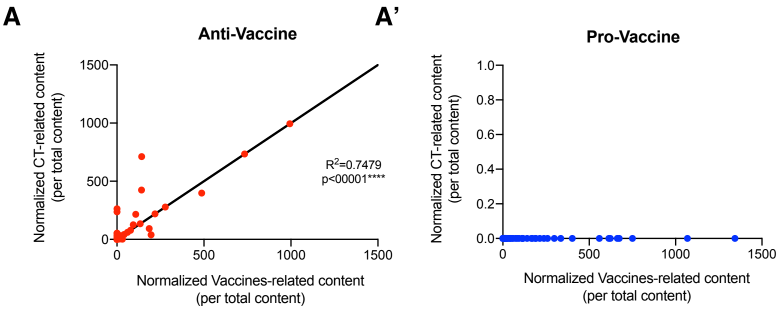


**Supplementary Figure 4. Positive correlation between the number of normalized vaccines-related contents and conspiracy theories-associated for the anti-vaccination group.** For both the anti-vaccination group (red), the higher the normalized number of vaccines-related contents generated or shared (for the overall number of contents generated on any given topic), the larger the number of normalized conspiracy theory (CT)-related tweets and retweets ($R^2$=0.7479; ****p<0.0001) (**A**). For the pro-vaccination group, no correlation exists between the normalized number of vaccines-related tweets and the normalized number of tweets and retweets including CTs (**B**). Outliers were excluded with ROUT, Q=0.1%; n=50.

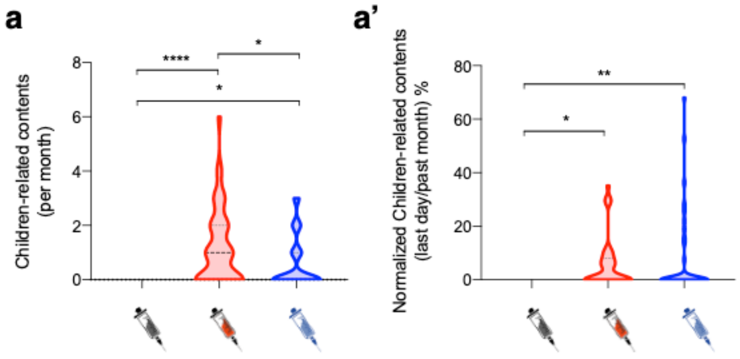


**Supplementary Figure 5. Both anti- and pro-vaccination groups share contents associated to children, but with different strategy and language.** Anti- (red) and pro-vaccination (blue) profiles share children-related contents, with the anti-vaccination group being the largest net producer of children-related contents on Twitter (**A**). We calculated the number of children-related content (tweets and retweets) published in the 24 hours before data analysis and normalized it for the total number of tweets published on average during a single day. 100 percent indicates that all generated contents are estimated to be children-related. Natural fluctuations above 100 percent are due to the variation between the activity on Twitter during the 24 hours prior to data analysis compared to an average day (**A'**). Representative examples of children-related tweets in the pro-vaccination group (**B**) and anti-vaccination group (**B'**). Representative examples of tweets concerning children, typical of the anti-vaccination group (**B''**). Ordinary one-way ANOVA; *p<0.05; **p<0.01;****p<0.0001; Outliers were excluded with ROUT, Q=0.1%; n=50.

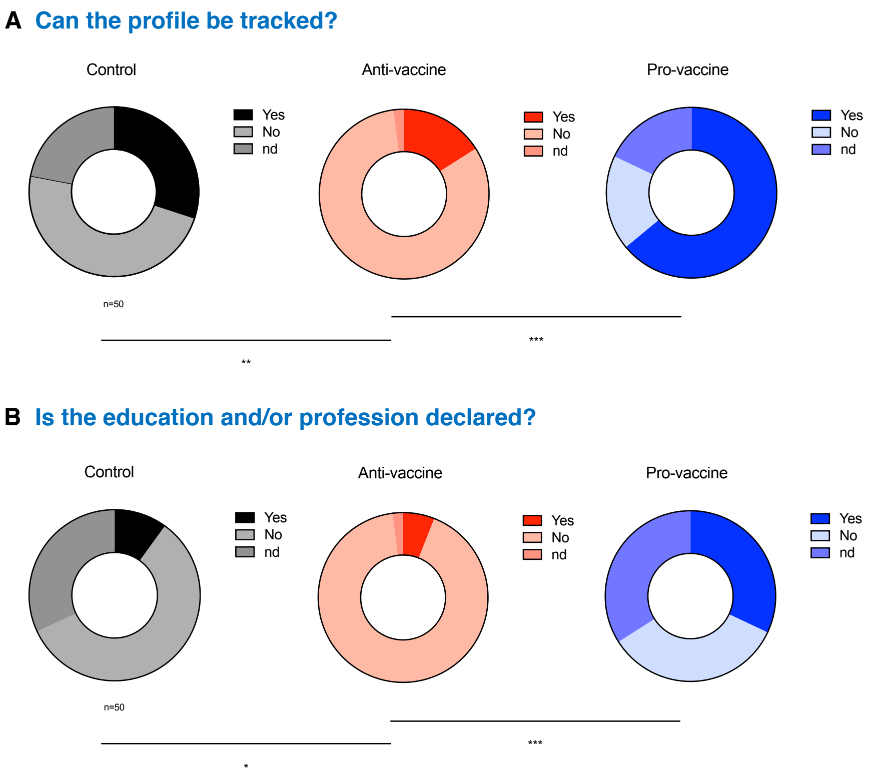


**Supplementary Figure 6. Anti-vaccination profiles are less prone to declare their identity, education or profession when compared with control and pro-vaccination profiles.** 30% of control profiles (grey shades) declare their identity (name and surname, and a profile picture depicting a real person). In comparison, pro-vaccination profiles (blue shades) are more likely declare their identity (64%) and only 16% of anti-vaccination profiles (red shades) declare their identity (**A**). 10% of control profiles declare either their education level or current profession. This percentage increases substantially for the pro-vaccination group (32%) and drops further for the anti-vaccination group (6%) (**B**). Profiles are defined as trackable when the user publicly releases her name, surname and a valid profile picture. Profiles are not defined as trackable when they fail to meet one of the aforementioned parameters. nd (not defined) indicates the above-mentioned criteria are not applicable (for instance, in the case of institutions without a verified badge on Twitter). This approach was also used for determining whether users declare their education level or profession. Chi-square test; *p<0.05;**p<0.01;***p<0.001; n=50.

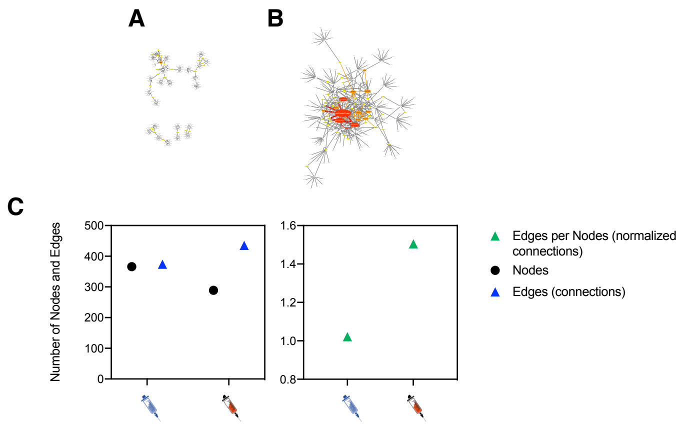


**Supplementary Figure 7. Anti-vaccination profiles are better connected with each other and establish a community, when compared with the pro-vaccination group.** The pro-vaccination (**A**) and anti-vaccination (**B**) Twitter webs, scaled 1:1. Yellow color represents Twitter profiles (nodes) with 2 to 4 anti-vaccination profiles preferentially retweeting their contents within the top 10 most retweeted users (edges; $2 \leq E \leq 4$; n=42). Orange nodes represent profiles with 5 to 9 edges ($5 \leq E \leq 9$; n=42), whereas red nodes indicate profiles with more than 10 connecting edges ($E \geq 10$; n=42). Size of the nodes is linearly scaled depending on the number of edges connecting the node (**A, B**). Number of nodes and edges for anti- (blue syringe) and pro-vaccination groups (red syringe). The anti-vaccination group has more edges than nodes, when compared with the pro-vaccination group. The number of edges per node is higher in the anti-vaccination web, when compared with the pro-vaccination web (**C**). Graphical representation and web parameters were generated with Cytoscape.
