## Supplementary figures and images for "The anti-vaccination infodemic on social media: a behavioral analysis"

### Anti-Vaccination network

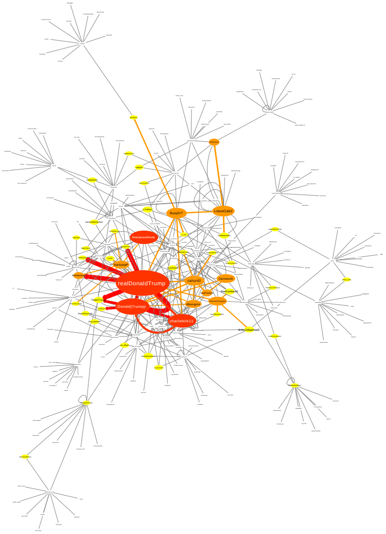

### Pro-vaccination network

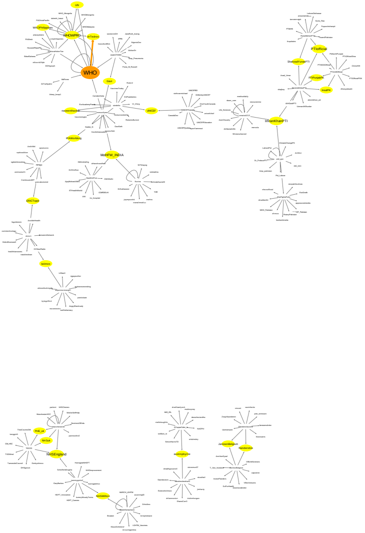
